# Fecal Biomarkers to Characterize Intestinal Inflammation among Children with Medically Attended Diarrhea across Six Low-Resource Settings: Findings from the Enterics for Global Health (EFGH) *Shigella* Surveillance Study

**DOI:** 10.64898/2026.05.01.26352214

**Authors:** Bri’Anna Horne, Brian Otieno Onyando, Henry Badji, Wardah Mujahid, Taufiqur Rahman Bhuiyan, Mutsai Bakali, Junaid Iqbal, Maribel Paredes Olortegui, Patricia Pavlinac, Bubacarr E. Ceesay, Olivia Schultes, Sharon M. Tennant, Billy Ogwel, Desiree Witte, Hannah Atlas, Naveed Ahmed, John Benjamin Ochieng, Khandra Sears, Sadia Islam, Queen Saidi, Samba Juma Jallow, Zarfishan Hussain, Paul Garcia Bardales, James A. Platts-Mills, Richard Omore, Farhana Khanam, Md. Parvej Mosharraf, Ousman Secka, Vitumbiko Munthali, Margaret N Kosek, Maureen Ndalama, Jen Cornick, Mohammad Tahir Yousafzai, M. Jahangir Hossain, Catherine Sonye, Firdausi Qadri, Elizabeth T. Rogawski McQuade, Stephanie A. Brennhofer

## Abstract

**Background:** Frequent enteric infections can damage the small intestine causing inflammation and malabsorption, leading to environmental enteric dysfunction. We aimed to characterize the association between intestinal inflammation and enteric pathogens among children in low- and middle-income countries (LMICs) who presented to care with diarrhea.

**Methodology/Principle Findings:** We conducted a cross-sectional analysis within the Enterics for Global Health - *Shigella* Surveillance Study at six LMIC sites: Bangladesh, Kenya, Malawi, Pakistan, Peru, and The Gambia. From August 2022 to July 2024, rectal swabs and whole stool samples were collected from 4,903 children with medically attended diarrhea aged 6-35 months (44.4% females, n=2178/4903; mean age: 15.4 months ± 7.4 months) and were analyzed for *Shigella* and four fecal inflammatory biomarkers: hemoglobin, lipocalin-2, myeloperoxidase, and calprotectin via Enzyme Linked Immunosorbent Assays. Caregivers and clinicians demonstrated moderate accuracy in identifying blood in stool compared to fecal hemoglobin (area under the curve (AUC)=0.70). Among 10 pathogens evaluated, *Shigella*-attributable diarrhea had the highest concentrations of calprotectin, hemoglobin, and myeloperoxidase. *Shigella* culture-/PCR+ episodes had intermediate levels of inflammation between culture-/PCR- and culture+ episodes. In multivariable models restricted to *Shigella* episodes, dysentery was positively associated and vomiting was negatively associated with biomarker concentrations, with the strongest associations observed for hemoglobin (dysentery geometric mean ratio: 14.91 μg/g (95% CI: 8.77, 25.36) and vomiting geometric mean ratio: 0.44 μg/g (95% CI: 0.24, 0.81)). Age, sex, and both acute and chronic malnutrition were not associated with inflammatory biomarker concentrations.

**Conclusions/Significance:** Hemoglobin appeared to be a more sensitive marker of blood in stool than visual observation. While *Shigella* was associated with heightened levels of all inflammatory biomarkers, hemoglobin was most strongly associated with *Shigella*, especially among attributable and culture positive episodes. The distinct clinical characteristics of *Shigella* were most closely associated with elevated hemoglobin concentrations, suggesting its potential utility as a point-of-care diagnostic.

**AUTHOR SUMMARY:** Diarrhea is common among children under five years of age in low- and middle-income countries (LMICs). Repeated diarrheal illnesses can damage the gut, leading to issues with growth and brain development. We examined fecal samples collected from 4,903 children with diarrhea who were enrolled in the Enterics for Global Health - *Shigella* Surveillance Study. We tested samples for diarrheal pathogens and measured four inflammatory biomarkers. We found that hemoglobin better identified blood in stool than visual observation of blood. Additionally, certain biomarkers (calprotectin, myeloperoxidase, and most notably hemoglobin) were higher when diarrhea was caused by bacteria such as *Shigella* than when diarrhea was caused by viruses. Also, we discovered that *Shigella* episodes identified using molecular diagnostics caused a similar illness as those identified by culture. These results support that *Shigella* diarrhea episodes identified by molecular diagnostics or culture are more inflammatory than other episodes of diarrhea and may require appropriate antibiotic treatment.

## INTRODUCTION

Enteric infections are highly prevalent in low- and middle-income countries (LMICs) where poor sanitary conditions can result in fecally contaminated environments [1,2]. Frequent or persistent enteric infections, especially with *Shigella*, can lead to intestinal inflammation, increased intestinal permeability, and bacterial translocation of luminal microbes, all of which activate innate and acquired immune responses [1,3]. The combination of these precursor events can result in environmental enteric dysfunction (EED), a largely subclinical condition characterized by villus atrophy, crypt hyperplasia, and accumulation of immune cells in the intestinal mucosa [1,3]. EED often goes undetected as it rarely causes acute gastrointestinal symptoms that prompt medical care, however its consequences are substantial, including impaired nutrient absorption and increased vulnerability to adverse child health outcomes [2].

There is a need to characterize inflammation in children during etiology specific diarrhea episodes to understand how these episodes may contribute to EED. Enteric inflammation has been linked with growth faltering [4], micronutrient deficiencies [5], and impaired neurodevelopment in early childhood in some settings [6]. Importantly, not all enteric pathogens contribute equally to intestinal inflammation [7]. For instance, *Shigella* is an inflammatory bacterial pathogen [8] that causes epithelial shedding, crypt hyperplasia, and immune cell infiltration [9]. While EED is confirmed histologically via a biopsy of the small intestine, this is not feasible in resource constrained settings or ethically viable for children where biopsy is not medically indicated [10]. Therefore, non-invasive techniques are needed to characterize intestinal damage in children in low resource settings.

Fecal biomarkers such as hemoglobin, lipocalin-2, myeloperoxidase, and calprotectin are non-invasive biomarkers used to characterize intestinal inflammation (hemoglobin, lipocalin-2, myeloperoxidase, calprotectin), neutrophil damage (myeloperoxidase, calprotectin), and enterocyte damage (lipocalin-2) [7]. Previous studies have demonstrated that fecal calprotectin [11,12], myeloperoxidase [12], and fecal occult blood [12] were higher in bacterial gastroenteritis compared to viral gastroenteritis. However, few studies have quantitatively evaluated whether these inflammatory markers can help distinguish the intestinal impact of specific pathogens or clarify the clinical significance of infections detected by culture versus molecular methods [7]. Prior work has demonstrated that *Shigella* culture-negative but quantitative Polymerase Chain Reaction (qPCR)-attributable cases represent true *Shigella* infections and should be managed as culture positive cases [13]. We propose inflammatory biomarkers could be used as independent predictors of etiology to evaluate whether culture-negative, but qPCR-attributable cases cause the same amount of intestinal damage as culture-positive cases.

This study aimed to characterize intestinal inflammation among children who presented to care with diarrhea in LMICs. We sought to 1) characterize the distribution and correlation of four inflammatory biomarkers by day of presentation and etiology, 2) compare detection of hemoglobin with reported blood in stool, 3) evaluate the association between *Shigella* detection by qPCR and culture with intestinal inflammation; and 4) assess how clinical risk factors for *Shigella* such as dysentery, fever, and vomiting are associated with biomarker levels. Studying these inflammatory responses could help identify a biomarker that could be used as a point-of-care diagnostic to guide clinicians on antibiotic treatment for shigellosis.

## METHODS

### Study Design and Population

We conducted a cross-sectional nested sub-study on intestinal inflammation among children who presented to care with diarrhea within the Enterics for Global Health (EFGH) - *Shigella* Surveillance Study. This sub-study was conducted at six sites: Bangladesh, Kenya, Malawi, Pakistan, Peru, and The Gambia. Primarily, the EFGH study utilized a facility-based enrollment of diarrhea cases, a three month follow-up period, and home stool collection within 24 hours for children who did not produce whole stool at the facility during enrollment [14]. Diarrhea was defined as 3 or more loose or watery stools in the previous 24 hours and the onset of illness was within 7 days of study enrollment. Follow-up data was used for this sub-study.

### Clinical Data and Specimen Collection

We collected information on child demographics (e.g., sex, age), anthropometric measurements (mid-upper arm circumference, weight, and length/height), and illness (e.g., dysentery, fever, vomiting) during enrollment. Additionally, we asked caregivers to fill out a diarrhea diary to track illness symptoms (e.g., number of loose stools, dysentery, vomiting, fever) and medication usage for 14 days.

During enrollment, three rectal swabs were collected from each participant using Nylon flocked swabs (Copan Diagnostics). The first swab (FLOQswab™) was placed in a dry tube (2-mL screw top vial compatible with a bead beater machine) and stored at −80°C for qPCR testing by TaqMan Array Card (TAC) for identification of enteric pathogens. The remaining two swabs were placed directly in Cary-Blair (CB) transport media (FecalSwab, 4C028S) and modified buffered glycerol saline (mBGS) (FecalSwab, mBGS made locally) respectively for *Shigella* species identification by culture [15]. During the study period, whole stool samples were collected at enrollment in sterile stool cup containers for the inflammatory biomarker sub-study, shipped to the laboratory and aliquoted. To increase sub-study sample size, home stool collections were conducted within 24 hours of enrollment for participants who did not produce a stool during their enrollment visit.

### Inflammatory Biomarkers

Whole stool samples were tested for four inflammatory biomarkers: hemoglobin [16], lipocalin-2 [17], myeloperoxidase [18], and calprotectin [19] using commercial kits. Stored stool samples were thawed and analyzed for each biomarker according to the manufacturer’s instructions. Up to 80 samples were tested per plate with duplicates of standards and controls for each plate. The results were uploaded to a custom-built R-based shiny app, to derive a sigmoid 4-parameter curve from the standards to calculate concentrations for the samples. Samples with concentrations higher than the highest standard were further diluted and retested. Samples that remained higher than the highest standard after further dilution were imputed with the value of the highest standard multiplied by the highest dilution factor utilized for that sample. Samples that were lower than the lowest standard were assigned the value of the lowest standard divided by 2.

### Culture and qPCR for pathogens

Specimen testing was performed as part of the parent EFGH study. Methods are briefly described here. Rectal swabs were transported in two media: Cary-Blair (CB) and modified Buffer Glycerol Saline (mBGS) for culture on MacConkey and Xylose Lysine Decarboxylase agar. Each swab was cultured on two plates and incubated for 18-24 hours. Suspected *Shigella* colonies were subjected to biochemical testing on Triple Sugar Iron agar, Motility Indole Ornithine agar, and Urea medium, and incubated for 24 hours. *Shigella* was confirmed through latex agglutination targeting unique lipopolysaccharides found on the surface (antigens) of bacteria and the isolates were stored in Tryptic Soy Broth+15% glycerol [15]. Total nucleic acid was extracted from the dry swabs or whole stool if dry swabs were unavailable, for the detection of enteric pathogens using qPCR through the TAC platform. Pathogens were included in the analysis if the prevalence was ≥5% in the samples at Ct < 35 and could be attributed as the cause of diarrhea. Pathogens were attributed as the cause of diarrhea if detected at quantities higher (Ct lower) than the EFGH pathogen-specific quantity cutoff for attribution of diarrhea [20]. Episodes could be attributed to multiple pathogens. Separate cut offs were used for whole stools and rectal swabs as previously [20]. For each pathogen, the Ct value was transformed into log_10_ copies (35-Ct value/3.322).

### Data Analysis

#### Correlation between biomarkers

We evaluated the correlation between biomarkers (hemoglobin, lipocalin-2, myeloperoxidase, and calprotectin) using log_10_ concentrations overall and by individual sites. We also compared biomarker concentration by day of presentation to care relative to illness onset.

#### Comparison of hemoglobin and report of blood

We compared the log_10_ concentrations of hemoglobin in all collected stools with and without reported visible blood (as indicated by the caregiver or clinician at the screening, enrollment, or discharge visits or noted in the diarrhea diary). A receiver operating characteristics (ROC) curve analysis was conducted to evaluate how well reported visible blood in stools corresponded with laboratory detected blood in stools (i.e., log_10_ concentration of hemoglobin) from the enrollment samples. An optimal cut-point was identified using Youden’s index to quantify the sensitivity and specificity of reported visible blood in stool to identify elevated hemoglobin concentrations.

#### Distribution of biomarkers by pathogens

We characterized the distribution of biomarker log_10_ concentrations across all samples for each of the 10 analytic pathogens (*Shigella*, typical enteropathogenic *Escherichia coli* (tEPEC), *Campylobacter jejuni/coli*, heat-stable enterotoxigenic *E. coli* (ST-ETEC), *Cryptosporidium*, adenovirus 40/41, astrovirus, norovirus GII, rotavirus, and sapovirus). Analyses were further restricted to watery diarrhea episodes only. Furthermore, we calculated the mean and standard deviation of the log_10_ concentrations of the four biomarkers by pathogen. We also performed pairwise correlations among log_10_ biomarkers by pathogen. To evaluate the relationship between pathogen log_10_ copies and biomarker concentration, we ran a linear regression with generalized estimating equations (GEE) to account for repeated enrollments and adjusted for country to account for site differences. We also performed a sensitivity analysis using linear regression restricted to one enrollment per child where the first instance of *Shigella* in a stool sample was included or in the instance that no *Shigella* was present in any stool sample, the stool sample from the child’s first enrollment was included.

#### Distribution of biomarkers by Shigella diagnostic results

We assessed the distribution of log_10_ biomarker concentrations for *Shigella* by three categories: *Shigella* culture-/*Shigella* qPCR-, *Shigella* culture-/*Shigella* qPCR+, and *Shigella* culture+ regardless of qPCR result. We further evaluated differences in biomarker concentrations by qPCR result where the diarrhea episode was attributed to *Shigella* (whole stool: Ct < 29.8, rectal swab: Ct <29.5), where *Shigella* was detected but not attributed as the etiology (Ct ≥ 29.8 (whole stool) or ≥29.5 (rectal swab) but < 35), and where *Shigella* was not detected (Ct ≥ 35).

#### Risk factors for shigellosis

To determine whether demographic, anthropometric, or illness risk factors were associated with intestinal inflammation among *Shigella*-positive children, we ran a multivariable linear regression with GEE to account for repeated measures and adjusted for country to account for site differences.

All analyses were performed via R software version 4.0.2.

### Ethical Considerations

This analysis involves data collected as part of the EFGH protocol which was reviewed and approved by the Institutional Review Boards (IRB) and the Scientific and Ethical Review Committees of the EFGH collaborating institutions at the study sites. Written informed consent was obtained from the parent(s) or primary caregiver(s) of each child who met eligibility criteria before any research activities were performed.

## RESULTS

From August 2022 to July 2024, a total of 4,903 children aged 6–35 months from the six study sites with whole stools collected within 24 hours of enrollment were included in this study. The mean (SD) biomarker concentrations were 0.04 μg/g (SD: 167.67, n = 4903) for hemoglobin, 14.33 μg/g (SD: 10.33, n = 4903) for lipocalin-2, 1232.53 ng/mL (SD: 5.42, n = 4901) for myeloperoxidase, and 81,977 ng/mL (SD: 7.71, n = 4903) for calprotectin.

### Correlation between biomarkers

Myeloperoxidase and calprotectin were moderately correlated (*R* = 0.70), whereas correlations among the other biomarkers were weak, ranging from *R* = 0.31 to *R* = 0.44 **(Figure 1)**. Myeloperoxidase and calprotectin were moderately correlated across all sites (*R* = 0.70-0.74) except Bangladesh (*R* = 0.55) **(Supplemental Figure 1)**. Hemoglobin and myeloperoxidase (*R* = 0.25-0.36) and hemoglobin and lipocalin-2 (*R* = 0.23-0.38) had the weakest correlations across sites.

**Figure 1.**
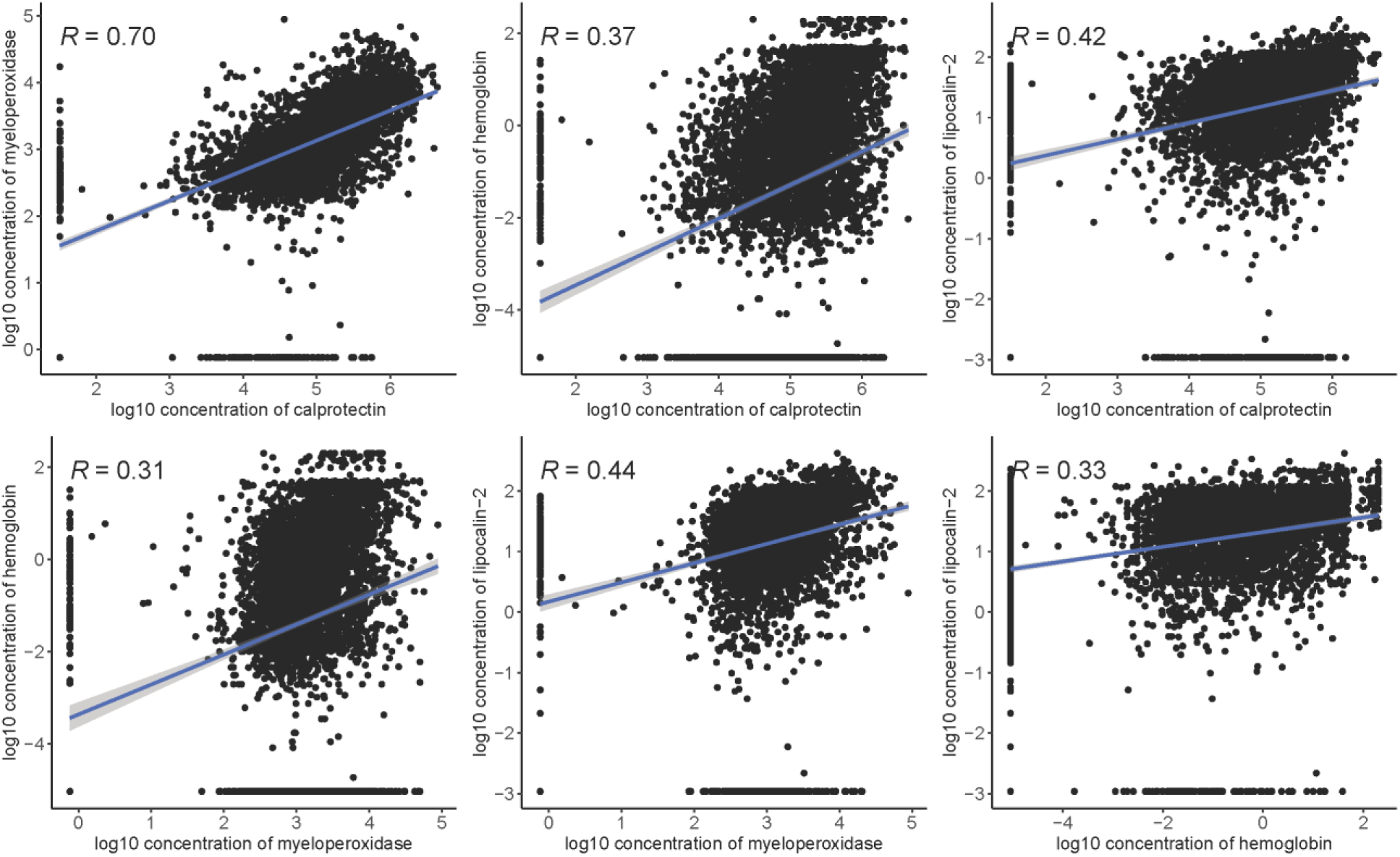
Pairwise correlations among log10 biomarker concentrations among children with medically attended diarrhea across the six sites in the EFGH sub-study.

### Comparison of hemoglobin and report of blood

Among all episodes, hemoglobin concentrations displayed a bimodal distribution when stratified by reported visible blood in stool, suggesting overall agreement between visible and ELISA detected blood, but with some notable discrepancies (**Figure 2**). There was more concordance among reported visible blood in stool and hemoglobin concentrations in *Shigella*-attributable diarrhea (**Supplemental Figure 2**) than *Shigella* detected but not attributed (**Supplemental Figure 3**) and *Shigella* not detected (**Supplemental Figure 4**). Overall, caregivers and clinicians were fairly accurate in predicting blood in a child’s stool (AUC=0.70). At the optimal quantitative hemoglobin cut-point, sensitivity of reported blood was low, but specificity was high. Among children with elevated hemoglobin in stool, only 32% of caregivers and clinicians reported blood in stool (**Supplemental Table 1**). Additionally, among children without hemoglobin in the stool, 91% of caregivers and clinicians confirmed there was no blood in the stool.

**Figure 2.**
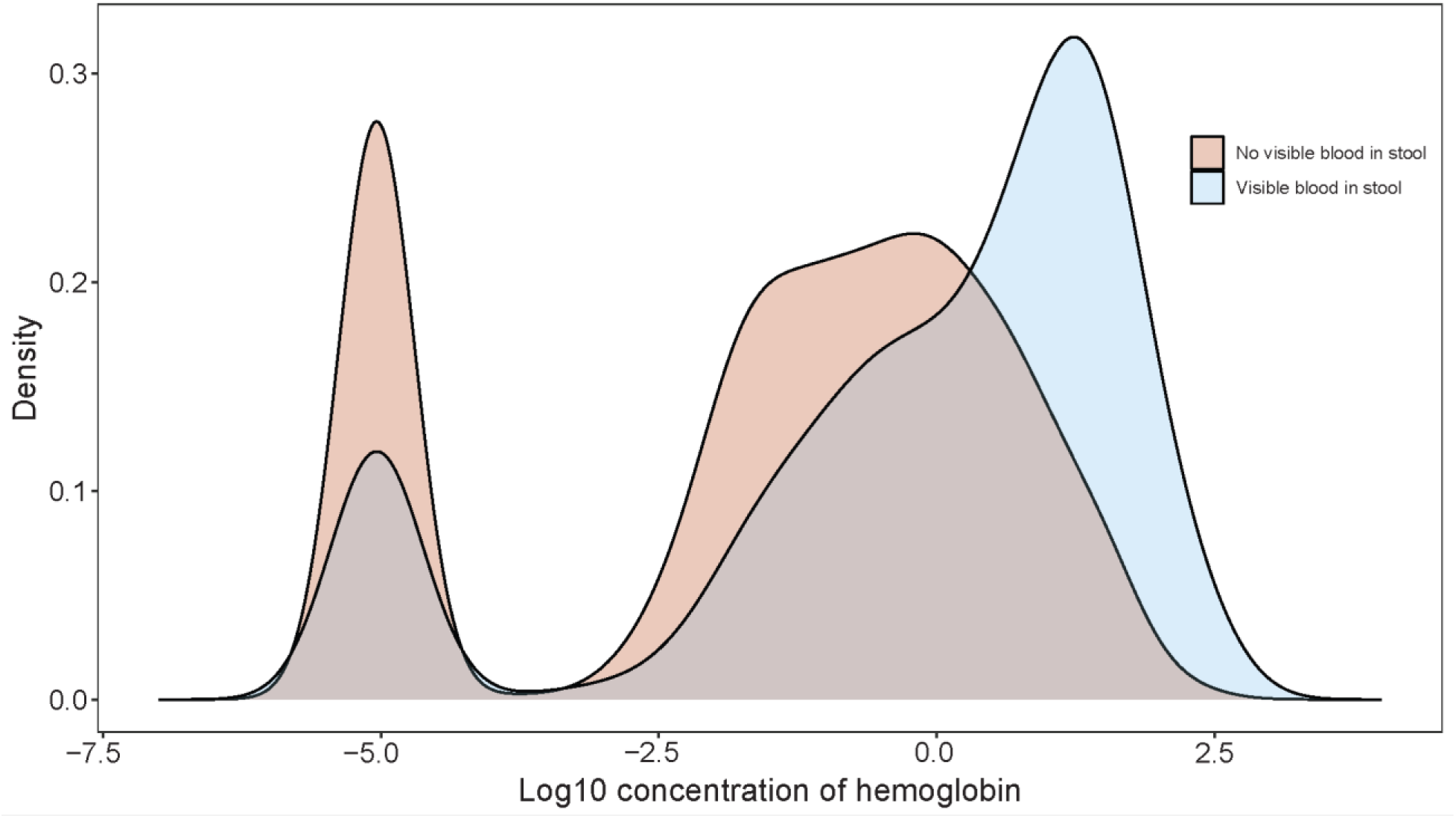
Density plot characterizing the distribution of log_10_ hemoglobin concentrations by caregiver and clinician reported visible blood in stool at screening, enrollment or discharge visits or noted in the diarrhea diary among children with medically attended diarrhea across the six sites in the EFGH sub-study.

Concentrations of biomarkers did not vary depending on day of presentation to care (hemoglobin: 4% higher concentration per day (95% CI: -6, 17); lipocalin-2: 5% higher concentration per day (95% CI: 0, 11); myeloperoxidase: 3% lower concentration per day (95% CI: -6, 1); calprotectin: 2% higher concentration per day (95% CI: -2, 6). In contrast, caregivers were 11% more likely to report visible blood in stool for each additional day of the diarrheal episode (RR: 1.11; 95% CI: 1.06–1.17), indicating that blood was more frequently observed later in the course of illness.

### Distribution of biomarkers by pathogens

Of the 10 analytic pathogens, *Shigella* (22.7%) had the highest and astrovirus (1.3%) had the lowest prevalence of attributable episodes (**Supplemental Table 2**). *Shigella* had the highest concentrations across all biomarkers (hemoglobin mean: 0.47 μg/g, SD: 118.36; myeloperoxidase mean: 2004.96 ng/mL, SD: 4.3; and calprotectin mean: 162588 ng/mL, SD: 5.3), except lipocalin-2 (mean: 21.62 μg/g, SD: 8.05) where *C. jejuni/coli* was associated with the highest concentration (mean: 25.78 μg/g, SD: 4.28) (**Figure 3; Supplemental Table 3**). Results were consistent when restricted to watery diarrhea (**Supplemental Figure 5**). *Shigella* was the only pathogen to have a positive association with all four biomarkers (**Supplemental Figure 6**). Of note, hemoglobin concentrations were markedly higher among *Shigella*-attributed episodes than non-attributed episodes (128% higher (95% CI: 102, 157), followed by calprotectin (26% higher; 95% CI: 20, 32), myeloperoxidase (20% higher; 95% CI: 15, 24), and lipocalin-2 (16% higher; 95% CI: 9, 23) (**Table 1**). The viral pathogens (astrovirus, norovirus GII, rotavirus, and sapovirus) had negative associations across all four biomarkers (**Supplemental Figure 6, Table 1).** A sensitivity analysis that restricted to one enrollment per child was consistent with the main findings (**Supplemental Table 4**).

**Figure 3.**
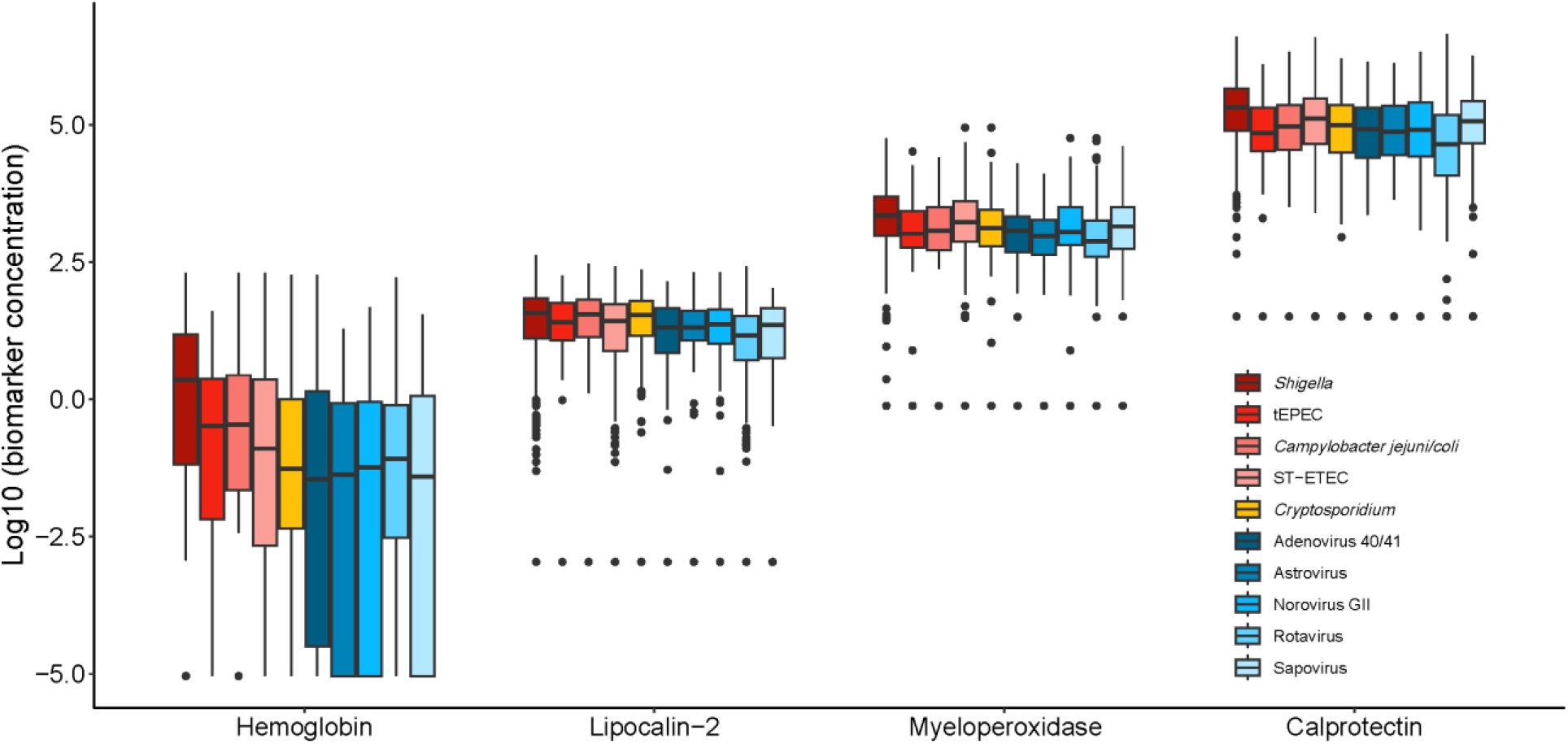
Boxplots characterizing the distribution of log10 biomarker concentrations by the 10 most prevalent enteric pathogens among children with medically attended diarrhea across the six sites in the EFGH sub-study.

**Table 1.** Associations of individual biomarkers with pathogen specific diarrhea etiologies among children with medically attended diarrhea across the six sites in the EFGH sub-study.

|  | Hemoglobin log<sub>10</sub><br>Mean Difference<br>(95% CI) | Lipocalin-2 log<sub>10</sub><br>Mean Difference<br>(95% CI) | Myeloperoxidase<br>log<sub>10</sub> Mean Difference<br>(95% CI) | Calprotectin log<sub>10</sub><br>Mean Difference<br>(95% CI) | Hemoglobin<br>Ratio<br>(95% CI) | Lipocalin-2<br>Ratio<br>(95% CI) | Myeloperoxidase<br>Ratio<br>(95% CI) | Calprotectin<br>Ratio<br>(95% CI) |
| --- | --- | --- | --- | --- | --- | --- | --- | --- |
| *Shigella* | 0.36 (0.30, 0.41) | 0.06 (0.04, 0.09) | 0.08 (0.06, 0.09) | 0.10 (0.08, 0.12) | 2.28 (2.02, 2.57) | 1.16 (1.09, 1.23) | 1.20 (1.15, 1.24) | 1.26 (1.20, 1.32) |
| tEPEC | 0.04 (-0.05, 0.13) | -0.02 (-0.05, 0.02) | 0.01 (-0.02, 0.03) | -0.01 (-0.04, 0.03) | 1.10 (0.90, 1.35) | 0.96 (0.89, 1.05) | 1.01 (0.95, 1.08) | 0.98 (0.91, 1.06) |
| *C. jejuni/coli* | 0.10 (0.03, 0.17) | 0.04 (0.02, 0.07) | -0.00 (-0.03, 0.02) | -0.01 (-0.03, 0.02) | 1.27 (1.08, 1.50) | 1.10 (1.04, 1.17) | 0.99 (0.94, 1.04) | 0.99 (0.93, 1.05) |
| ST-ETEC | -0.09 (-0.16, -0.01) | -0.02 (-0.06, 0.01) | 0.03 (0.01, 0.05) | 0.04 (0.01, 0.06) | 0.82 (0.70, 0.97) | 0.95 (0.88, 1.03) | 1.06 (1.02, 1.11) | 1.09 (1.03, 1.16) |
| *Cryptosporidium* | -0.08 (-0.18, 0.01) | 0.06 (0.02, 0.10) | -0.01 (-0.05, 0.02) | -0.03 (-0.08, 0.02) | 0.83 (0.67, 1.03) | 1.16 (1.06, 1.26) | 0.97 (0.90, 1.05) | 0.93 (0.84, 1.04) |
| Adenovirus 40/41 | -0.06 (-0.13, 0.01) | -0.00 (-0.03, 0.02) | -0.03 (-0.05, -0.01) | -0.04 (-0.08, -0.01) | 0.87 (0.74, 1.03) | 0.99 (0.93, 1.05) | 0.94 (0.89, 0.99) | 0.90 (0.84, 0.97) |
| Astrovirus | -0.30 (-0.57, -0.04) | -0.06 (-0.16, 0.05) | -0.09 (-0.17, 0.00) | -0.01 (-0.11, 0.10) | 0.50 (0.27, 0.91) | 0.88 (0.69, 1.11) | 0.82 (0.67, 1.00) | 0.99 (0.77, 1.26) |
| Norovirus GII | -0.28 (-0.44, -0.12) | -0.05 (-0.10, 0.00) | -0.03 (-0.08, 0.01) | -0.09 (-0.15, -0.03) | 0.53 (0.37, 0.76) | 0.89 (0.79, 1.01) | 0.92 (0.83, 1.03) | 0.81 (0.71, 0.93) |
| Rotavirus | -0.15 (-0.25, -0.05) | -0.05 (-0.08, -0.01) | -0.05 (-0.08, -0.01) | -0.10 (-0.14, -0.05) | 0.71 (0.56, 0.89) | 0.90 (0.82, 0.99) | 0.90 (0.83, 0.97) | 0.80 (0.72, 0.88) |
| Sapovirus | -0.16 (-0.30, -0.03) | -0.06 (-0.12, -0.00) | -0.04 (-0.08, 0.01) | -0.01 (-0.05, 0.04) | 0.69 (0.50, 0.93) | 0.87 (0.76, 1.00) | 0.92 (0.84, 1.01) | 0.99 (0.89, 1.10) |

tEPEC = typical enteropathogenic *Escherichia coli*; *C. jejuni/coli*: *Campylobacter jejuni/coli*; ST-ETEC = heat-stable enterotoxigenic *Escherichia coli*
Hemoglobin back-transformed units: µg/g; Lipocalin-2 back-transformed units: µg; Myeloperoxidase back-transformed units: ng/mL; Calprotectin back-transformed units: ng/mL.
Note: Ratios are of geometric means. Back-transformed (antilogged) values were calculated using unrounded log<sub>10</sub> estimates; therefore, small discrepancies may occur if rounded log<sub>10</sub> values are back-transformed directly.

### Distribution of biomarkers by Shigella diagnostic results

*Shigella* culture-/qPCR+ biomarker concentrations were statistically different from *Shigella* culture-/qPCR- episodes and *Shigella* culture+ regardless of qPCR result (**Figure 4**). *Shigella*-attributed episodes (**Figure 5**) and *Shigella* culture-positive episodes (**Supplemental Figure 7)** had higher concentrations of inflammatory biomarkers than non-attributed and culture-negative episodes, respectively.

**Figure 4.**
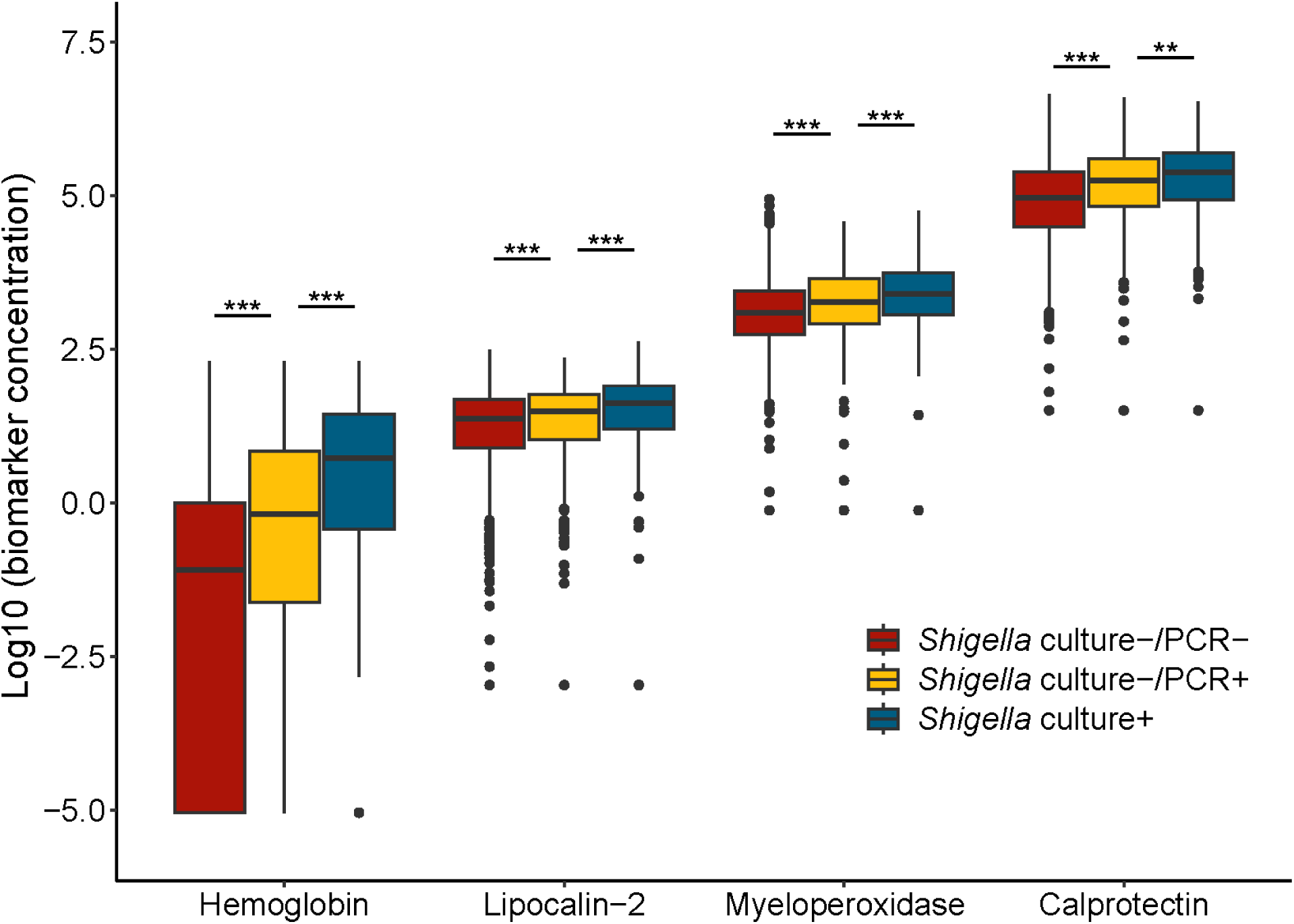
Boxplots characterizing the distribution of log_10_ biomarker concentrations by *Shigella* PCR and culture positivity status among diarrhea cases among children with medically attended diarrhea across the six sites in the EFGH sub-study. ** = p< 0.01; *** = p< 0.001

**Figure 5.**
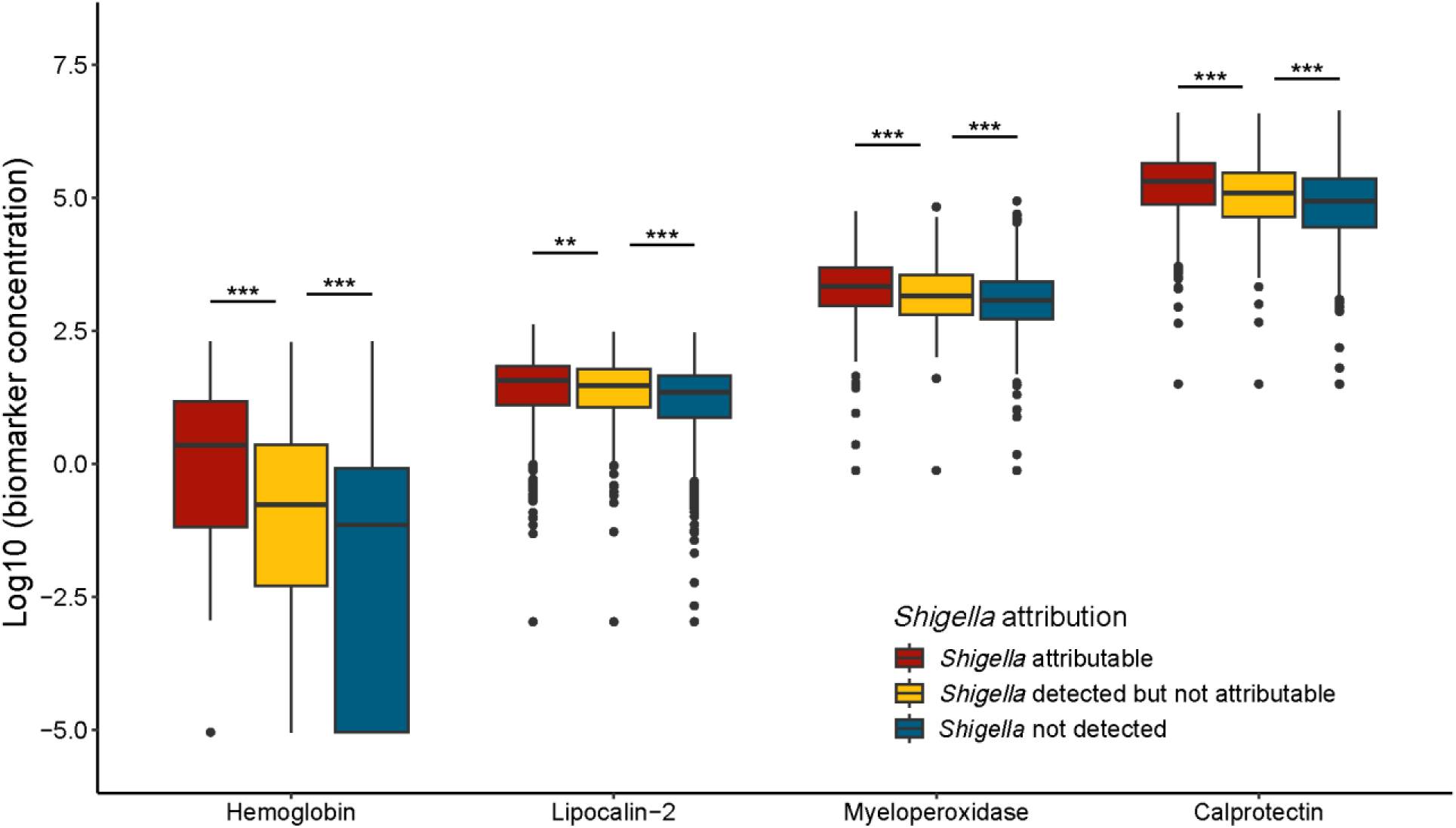
Boxplots characterizing the distribution of log10 biomarker concentrations by *Shigella* attribution status among diarrhea cases among children with medically attended diarrhea across the six sites in the EFGH sub-study. ** = p< 0.01; *** = p< 0.001

### Risk factors for shigellosis

Among *Shigella*-attributed episodes, dysentery was the strongest risk factor for inflammation across biomarkers but was most notable for hemoglobin where hemoglobin concentrations were 14 times higher among dysentery cases compared to non-dysentery cases (geometric mean ratio: 14.91 μg/g; 95% CI: 8.77, 25.36) (**Figure 6; Supplemental Table 5**). Additionally, having a fever was positively associated across inflammatory markers, significantly so with hemoglobin (geometric mean ratio: 1.89 μg/g; 95% CI: 1.05, 3.42). In contrast, vomiting was negatively associated with all inflammatory biomarkers, especially for hemoglobin (geometric mean ratio: 0.44 μg/g; 95% CI: 0.24, 0.81). There was no association between age, sex, and malnutrition with any inflammatory biomarker.

**Figure 6.**
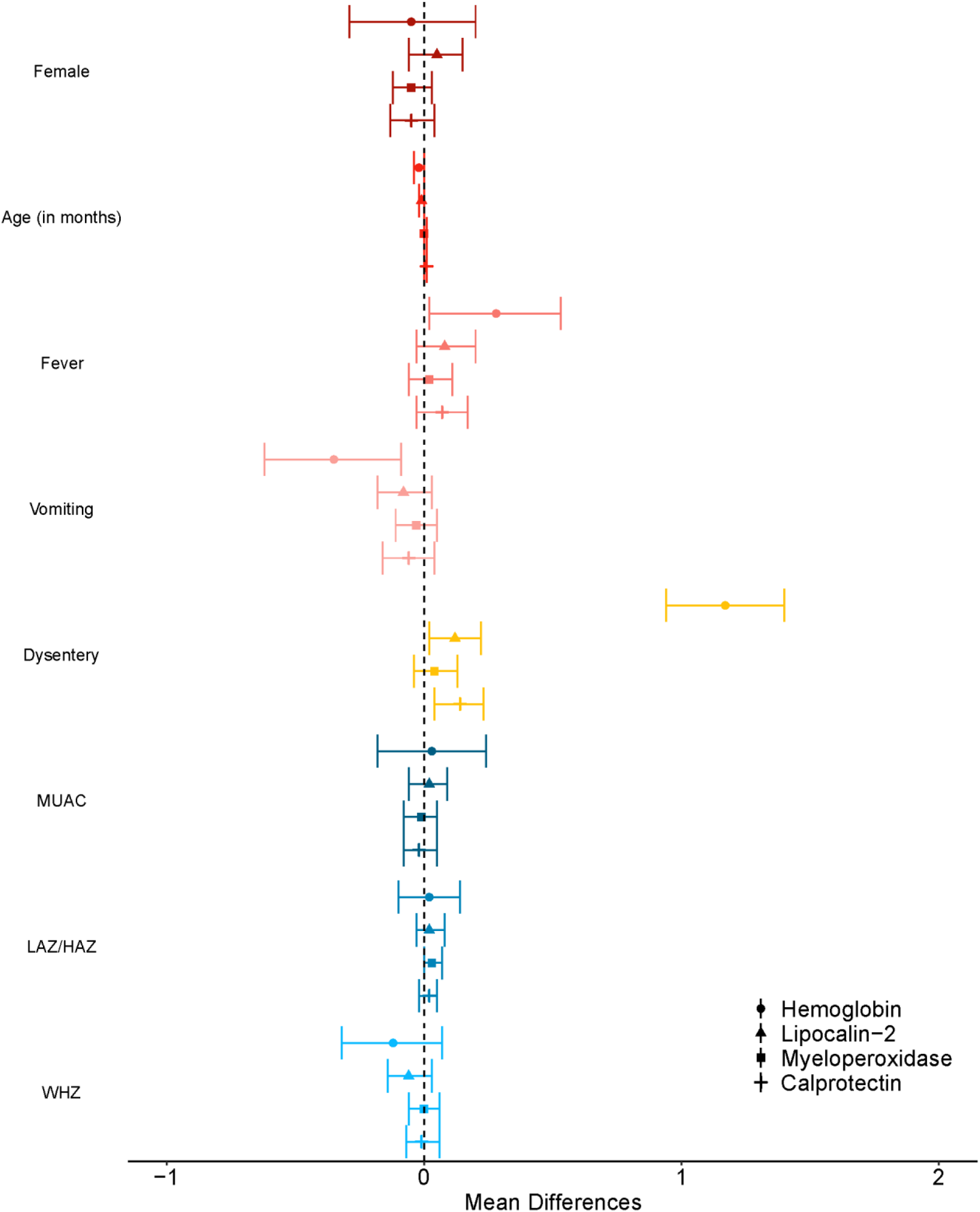
Associations of individual biomarkers with risk factors for shigellosis among children with medically attended diarrhea across the six sites in the EFGH sub-study. MUAC = mid-upper arm circumference; LAZ/HAZ = length for age z-score/height for age z-score; WHZ = weight for height z-score

## DISCUSSION

We conducted a large multisite site study on pediatric diarrhea to characterize intestinal inflammation by etiology. Diarrhea episodes with bacterial etiologies had higher concentrations of all four inflammatory biomarkers (hemoglobin, myeloperoxidase, lipocalin-2, and calprotectin) compared to episodes with viral or parasitic etiologies. These findings are consistent with previous pediatric studies which showed that fecal calprotectin [11,21], myeloperoxidase, [11] and fecal occult blood [11] were significantly higher in bacterial than viral gastroenteritis, with good diagnostic discrimination (AUCs ∼0.74–0.79) [7].

We observed a moderate correlation between myeloperoxidase and calprotectin, while weak correlations were observed among the other pairs, indicating that hemoglobin, myeloperoxidase and lipocalin-2 capture distinct elements of the inflammatory response. Myeloperoxidase and calprotectin may capture a similar response of neutrophil damage [7], which has been previously documented [12].

*Shigella*-attributable episodes had the highest concentrations of hemoglobin, myeloperoxidase, and calprotectin, while *Campylobacter jejuni/coli* had the highest concentrations of lipocalin-2. Among *Shigella* cases we observed the highest concentrations for each biomarker among the attributable cases, followed by the detected but not attributable cases, then the not detected cases. This would suggest that higher quantities of *Shigella* correspond to more inflammation. Furthermore, we found that culture-/qPCR+ *Shigella* episodes had elevated biomarker concentrations compared to culture-/qPCR-episodes, supporting that qPCR-attributed *Shigella* reflects clinically meaningful *Shigella* diarrhea rather than incidental carriage of *Shigella* during diarrhea. This would indicate that qPCR-attributable cases, even if culture negative, cause intestinal inflammation. This finding is consistent with prior work which demonstrated that *Shigella* culture-/qPCR+ cases represent true *Shigella* infections and should be managed clinically as culture+ cases would be managed [13].

In analyses restricted to *Shigella*-attributable episodes, dysentery was positively associated with all biomarkers, but the association was most pronounced for hemoglobin. This is unsurprising as hemoglobin is a marker of red blood cells [7]. Additionally, hemoglobin was also positively associated with fever and inversely associated with vomiting. These associations fit clinical expectations for *Shigella* diarrhea which is more likely to be associated with fever and bloody stools and less likely to be associated with vomiting [22] unless there is a co-etiology with a viral enteric pathogen [8]. Given hemoglobin’s close alignment with the presentation of *Shigella*, this could further underscore hemoglobin’s role as a biomarker for identifying diarrheal illnesses requiring antibiotic treatment.

The cross-sectional design of this study limited our ability to assess how biomarker levels changed over time. Children in LMICs tend to have high baseline levels of inflammation [23] so it is possible that we captured underlying enteropathy rather than inflammation due to the acute diarrhea illness for which the child was enrolled in the study. Nonetheless, our cross-sectional design was well suited for our aim of describing the effects of acute diarrheal illness. Future studies could implement longitudinal sampling particularly of fecal hemoglobin to help characterize dynamics of the biomarkers, define an optimal testing window, and identify diagnostic thresholds in this population. Stool samples were collected up to 24 hours after enrollment, at which time biomarker concentrations may have shifted. However, this window was necessary to allow for maximum stool collections across participants.

Hemoglobin showed promise in identifying shigellosis. It performed the best among the four biomarkers for distinguishing likely *Shigella* diarrhea, indicating that it may have an important role to play in identifying treatable diarrhea where advanced etiologic diagnostics are not available. Point-of-care studies testing for fecal occult blood show reasonable specificity for identifying invasive bacterial pathogens such as *Shigella* [7,25], which reinforces the utility of stool blood–based signals as part of an antibiotic decision framework in resource limited settings. Our results support further evaluation of hemoglobin to improve clinical prediction algorithms for shigellosis. Overall, our findings confirm the strong link between inflammatory stool markers and bacterial etiologies while establishing distinct pathogen-specific inflammatory signatures, providing a critical foundation for developing diagnostics to improve clinical decision-making.

## Data Availability

The EFGH statistical analysis plan (https://clinicaltrials.gov/study/NCT06047821) and study protocol (https://academic.oup.com/ofid/issue/11/Supplement_1) were made publicly available. The de-identified and anonymised dataset is published online (https://search.vivli.org/doiLanding/studies/PR00011860/isLanding) and analytic code is available at https://github.com/sb2yb/efgh-biomarker-substudy.

## ACKNOWLEDGEMENTS

This project is supported by the Bill & Melinda Gates Foundation (grant numbers: INV-016650, INV-031791, INV-036891, INV-036892, INV-028721, INV-041730, INV-044311, INV-044317).

